# Mutational analysis of SARS-CoV-2. ORF8 and the evolution of the Delta and Omicron variants

**DOI:** 10.1101/2021.12.19.21268069

**Authors:** Gopika Trieu, Vuong N Trieu

## Abstract

SARS-CoV-2 the virus responsible for the current pandemic. This virus is continually evolving, adapting to both innate and acquired immune responses and therapeutic drugs. Therefore, it is important to understand how the virus evolving to design the appropriate therapeutic and vaccine in preparation for future variants. Here, we used the online SARS-CoV-2 databases, Nextstrain and Ourworld, to map the evolution and epidemiology of the virus. We identified 30 high entropy residues which underwent a progressive evolution to arrive at the current dominant variant - Delta variant. The virus underwent mutational waves with the first wave made up of structural proteins important in its infectivity and the second wave made up of the ORFs important for its contagion. The most important driver of the second wave is ORF8 mutations at residue 119 and 120. Further mutations of these two residues are creating new clades that are offshoots from the Delta backbone. More importantly the further expansion of the S protein in the Omicron variant is now followed with the acquisition of ORF8 mutations 119 and 120. These findings demonstrate how SARS-CoV-2 mutates and points to two evolutionary paths; 1) Mutational expansion on the Delta backbone among the ORFs and 2) Mutational expansion of the S protein on other backbone follow with mutational wave among the ORFs. Both are happening at the same time right now with the Omicron variant early in the first wave to follow with a more aggressive second wave of mutations.

**HIGHLIGHTS:** Mutational waves in the evolution of SARS-CoV-2. S protein as the driver of the first wave improving the minimum inhaled viral load required to cause infection and ORF8 mutations 119 and 120 as the driver of the second mutational wave to improve the Contagion Airborne Transmission value.

## INTRODUCTION

Since late 2019, an outbreak of upper respiratory infection and pneumonia caused by a novel coronavirus (2019-nCoV) has rapidly spread from its epicenter in Wuhan in Hubei province, China to become a global epidemic with millions of cases and hundreds of thousands of deaths. The virus has now been named Severe Acute Respiratory Syndrome CoronaVirus 2 (SARS-CoV-2), and the disease it causes has been named CoronaVirus Disease 2019 (COVID 19). It is believed that the outbreak has a zoonotic origin, with animal-to-human transmission followed by human-to-human spread via aerosol droplets and contaminated surfaces. As with the prior outbreaks of Severe Acute Respiratory Syndrome (SARS) and Middle East Respiratory Syndrome (MERS), numerous approaches are being taken in an attempt to treat and prevent the disease. As of 12-15-2021 there are 271M infected and 5.32M dead worldwide. The highest affected country being the United States (50.3M infected and 801K dead), followed by India (34.7M infected and 476K dead), Brazil (22.2M infected and 617K dead), United Kingdom (10.9M infected and 147K dead), and Russia (9.9M infected and 286K). Except for India, all these countries are still struggling with the pandemic. This emphasizes how pervasive and recalcitrant the pandemic has become.

The genomic information for SARS-CoV-2 is known and has been shared. The SARS-CoV-2 genome encodes 28 confirmed proteins. Open reading frame 1ab (ORF1ab) encodes polyproteins PP1ab and PP1a which are cleaved into 16 nonstructural proteins (Nsp1 to Nsp16). Additionally, there are four structural proteins (spike [S], envelope [E], membrane [M], and nucleocapsid [N]) and eight accessory proteins (ORF3a, ORF3b, ORF6, ORF7a, ORF7b, ORF8, and ORF9b). Mutational conservation is highest for the Nsp polyproteins, while the genome sequence encoding the accessory factors (ORFs) diverges greatly [1]

Study of mutational changes of SARS-CoV-2 will help us understand the epidemiology of COVID-19 and project out potential mutational changes in the future so that interventional steps can be implemented to stop the pandemic.

## METHOD

Global SARS-CoV-2 genomic sequencing efforts have contributed large amounts of sequencing data from several variants into various public databases such as, GISAID and Nextstrain and NCBI SARS-CoV-2 Resources. These databases have allowed the interrogation of viral diversity with associated disease transmission in different countries [2]. In Nextstrain, the data is organized into a phylogeny tree showing evolutionary relationships of SARS-CoV-2 viruses. The site subsamples available genome data for these analysis views with ∼600 genomes per continental region (∼200 prior to the last 4 months, and ∼400 from the most recent 4 months) in order to display a balanced global sequence distribution. Site numbering and genome structure uses Wuhan-Hu-1/2019 as a reference and the phylogeny rooted relative to early samples from Wuhan. Temporal resolution assumes a nucleotide substitution rate of 8 × 10^−4^ substitutions per site per year. For each position, Nextstrain calculated the Shannon entropy of the distribution of amino acids [3] where a score of 0 corresponds to no variation and higher scores correspond to sites with increasing amino acid diversity.

Epidemiological data was obtained through https://ourworldindata.org/coronavirus [4].

## RESULTS

As shown below in Figure 1, there are high entropy residues across the SARS-CoV-2 genome. Residues with entropy greater than 0.6 were examined for their ability to generate offshoots from the Delta backbone. Each of the 30 high entropy residues examined was able to separate out Delta variants away from pre-Delta variants such as Alpha, Beta, and Omicron (Table 1). Of those, two are class 1 (S protein; residues 95 and 142) and exhibit random distribution across the Delta phylogeny tree. Four are class 2 (ORF1a; residue 2930, S protein; residue 158, ORF8; residues 119 and 120) and are start of new lineage on the Delta backbone. One is class 3 (ORF7a; residue 71) and showed a phylogenic linkage between two major branches of the Delta variant. The phylogeny trees displayed as divergence linked radial for these residues are shown below in Figure 2. As shown in Figure 2d, these mutational changes occur in waves. The first wave consists of the structural proteins with the N protein leading follow by the S protein. The second wave consists of ORFs proteins with ORF8 happening early.

**Table 1.**
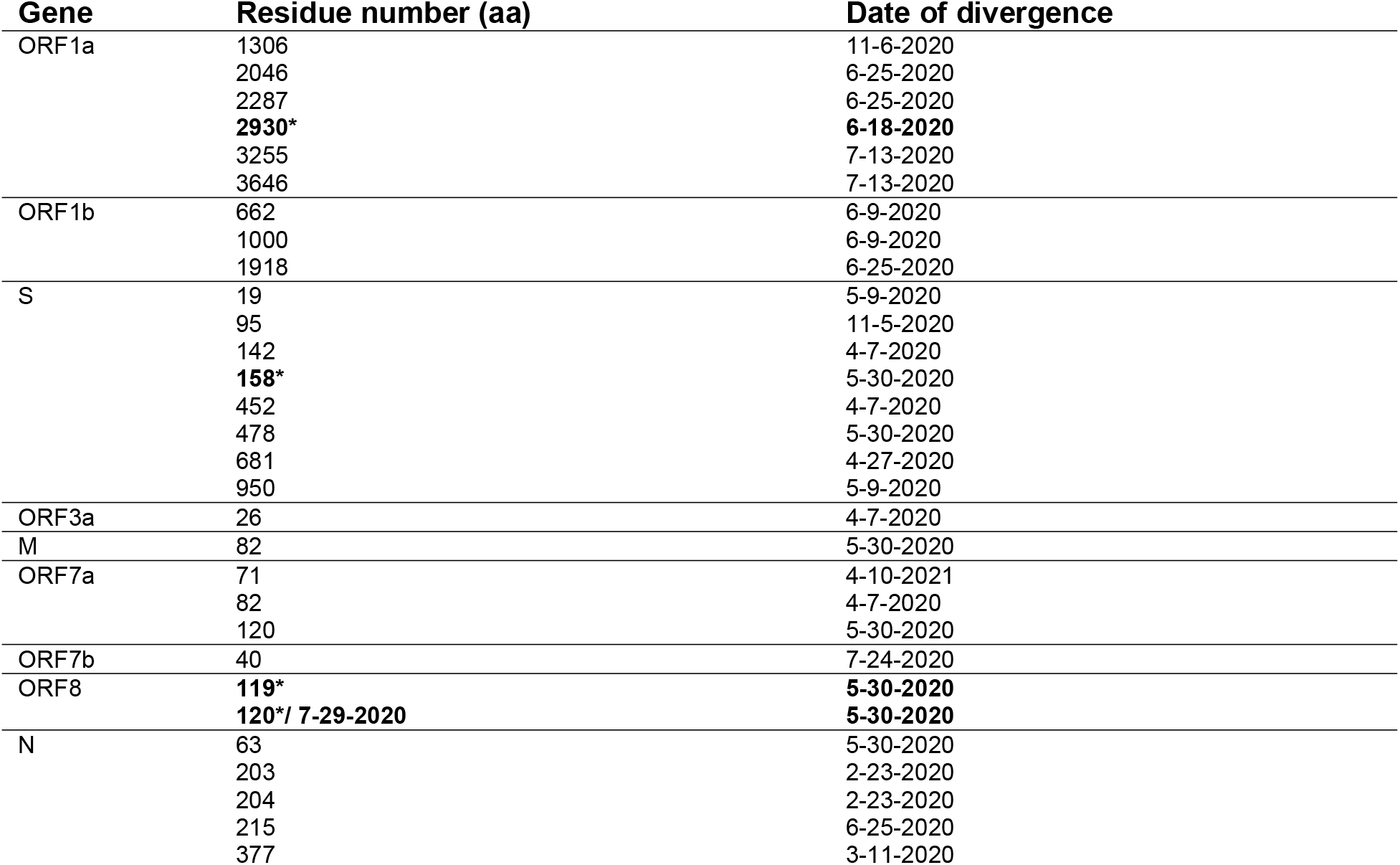
Hotspots examined for stable offshoots from delta backbone. Asterisk indicates residues capable of forming new offshoots.

**Figure 1a.**
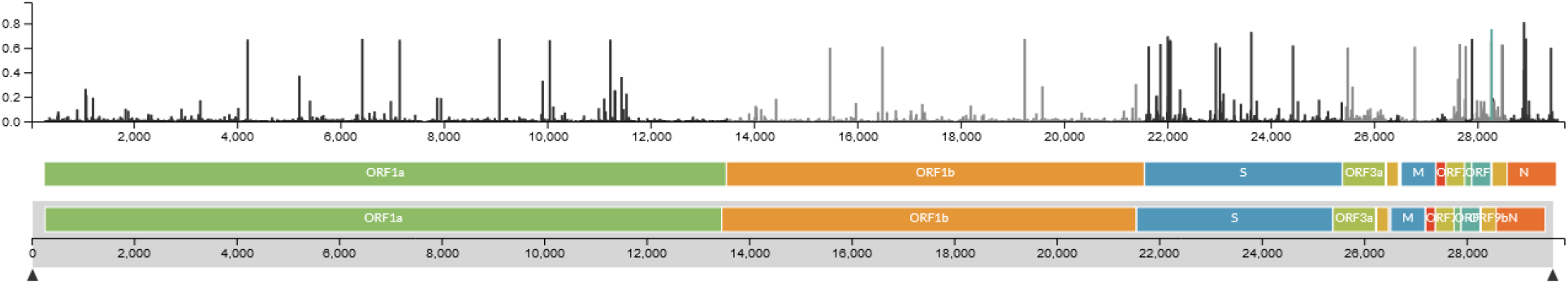
Mutational hotspots as measured by entropy score across the SARS-COV-2 genome

**Figure 2.**
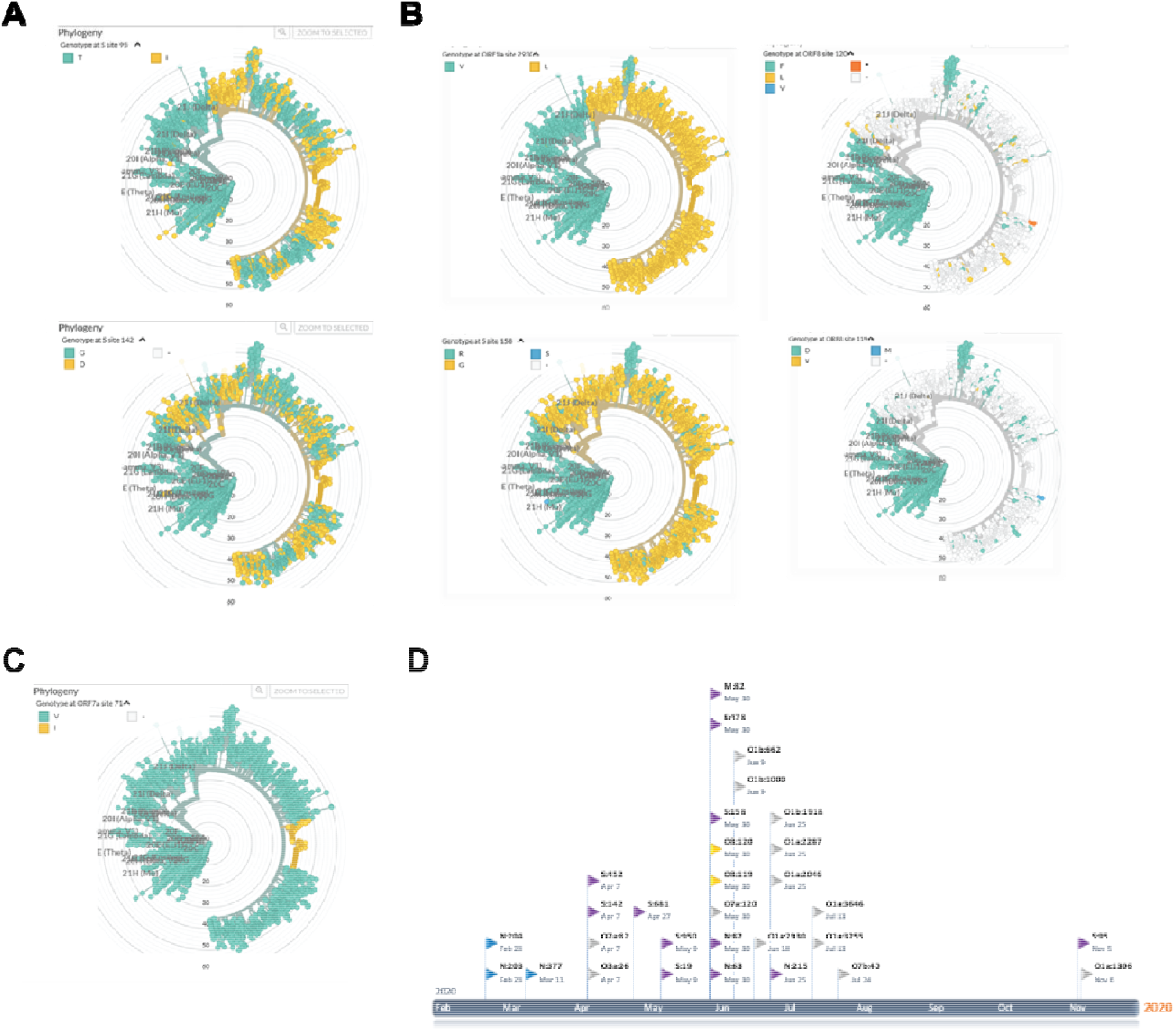
Mutational divergence analysis of SARS-CoV-2. A) Class 1 hotspots. Mix distribution B) Class 2 hotspots. New lineage C) Class 3 hotspots. Linkage Figure D) Mutational waves observed with SARS-CoV-2. Structural proteins are either purple (Spike protein) or blue (N and M proteins). Nonstructural ORF proteins are either in yellow (ORF8) or grey (all other ORFs). The amino acid residue and the date of divergence are indicated in text.

To further understand the role of ORF8 in the epidemiology of SARS-CoV-2, we scanned each residue of ORF8 for its ability to form new clades. As shown in Figure 3, only residues 27, 52, 73, 92, 119 and 120 were able to branch off into a new clade. Additionally, 52, 119 and 120 being part of the covalent dimer interface; 73 and 92 are part of the alternate dimeric interface.

**Figure 3.**
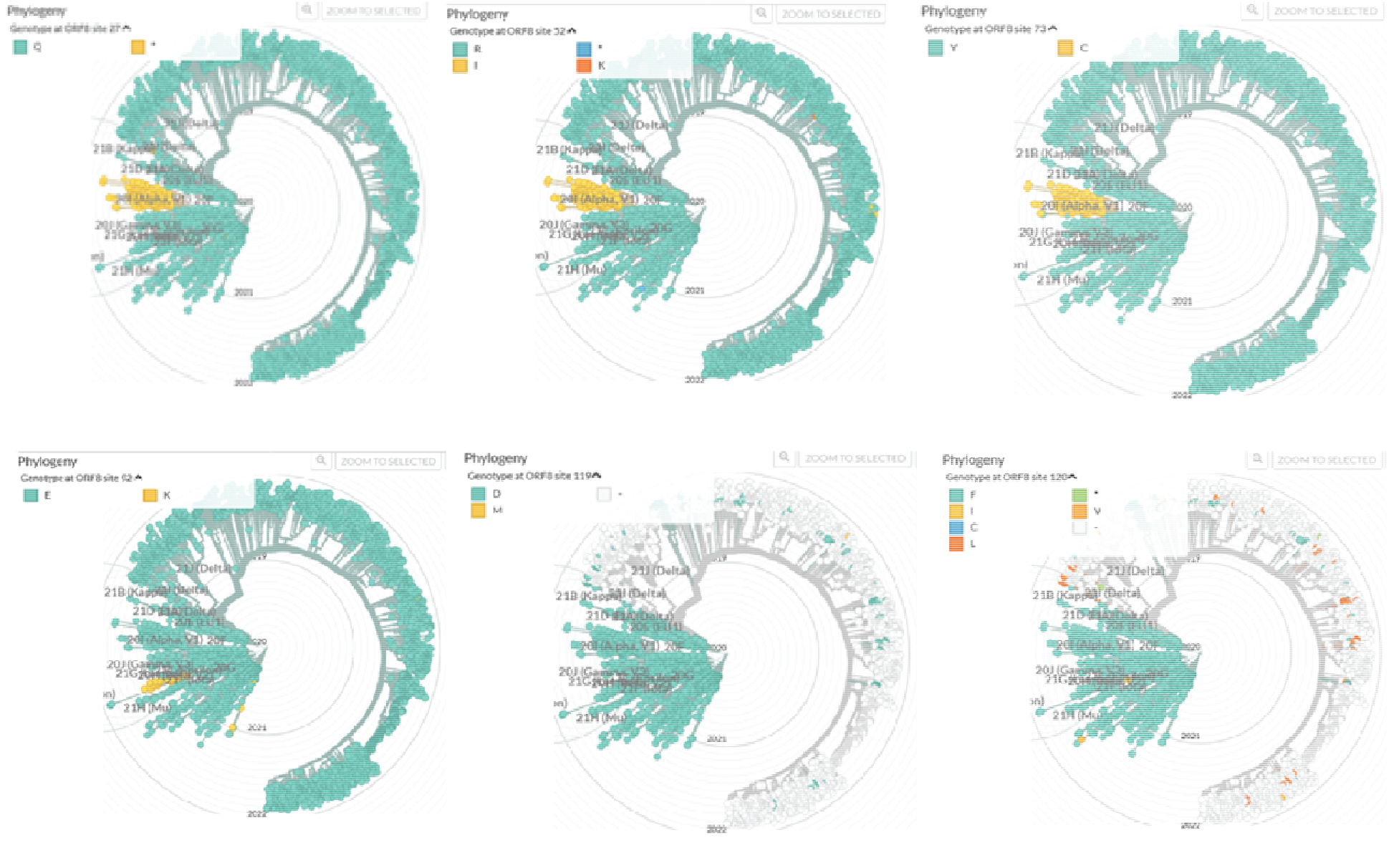
Formation of new clades by mutational changes in a single residue in ORF8

To determine if the 120 mutations have been disseminated globally, we performed the analysis with a regional subset of the data on Nextstrain from August 2021 to December 2021. The data shown in Figure 4 indicated that the mutations have spread widely into all regions under analysis with the dominant region being Egypt (F mutation) and India (L mutation). In Egypt, a number of new cases have been increasing beginning June 2021 but have now plateaued; case fatality rate has been fluctuating but there is no trend towards an increase in case fatality (Figure 5). The data would suggest that F120 is potentially more contagious but not more virulent. In contrast, the case fatality rate in India has been increasing to its highest level since the pandemic, despite decreasing numbers of new cases suggesting that the -L120, though endemic like the situation in Egypt, is more virulent.

**Figure 4.**
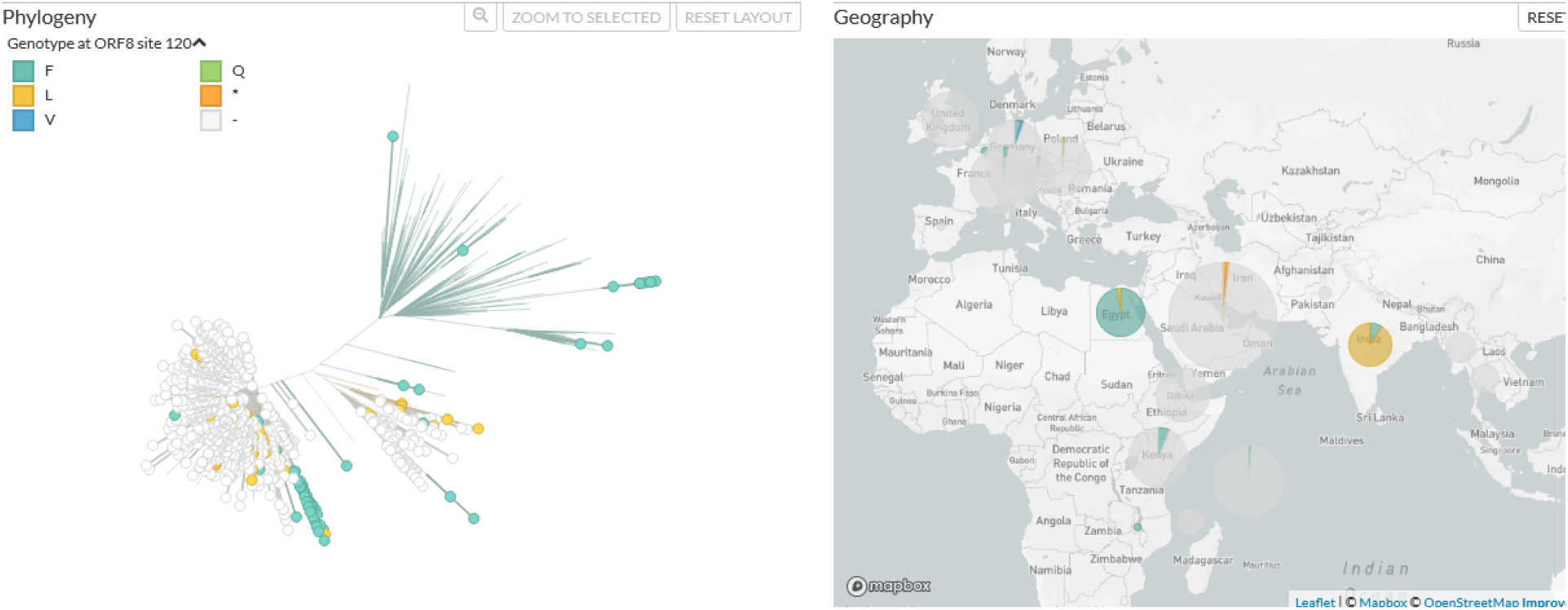
Phylogeny tree of residue 120 ORF8. The variants dominate Egypt for F mutation and L mutation for India.

**Figure 5.**
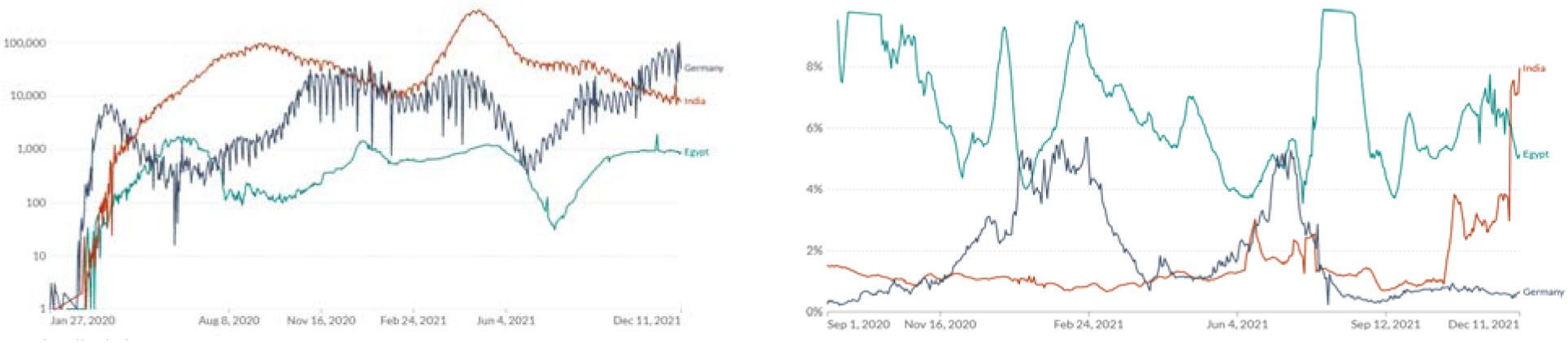
Confirmed cases and fatality rate of SARS-CoV-2. A) Daily confirmed cases of SARS-CoV-2 B) Moving average case fatality rate of SARS-CoV-2

We also examined the possibility that the emerging Omicron variant could acquire the F120-mutation characteristics of the Delta variant. As shown in Figure 6, this indeed is true. A fraction of the Omicron variants has acquired the F120-mutation. This variant should be monitored carefully so that it does not become widespread as the Delta variant once did following the completion of the second mutational wave..

**Figure 6.**
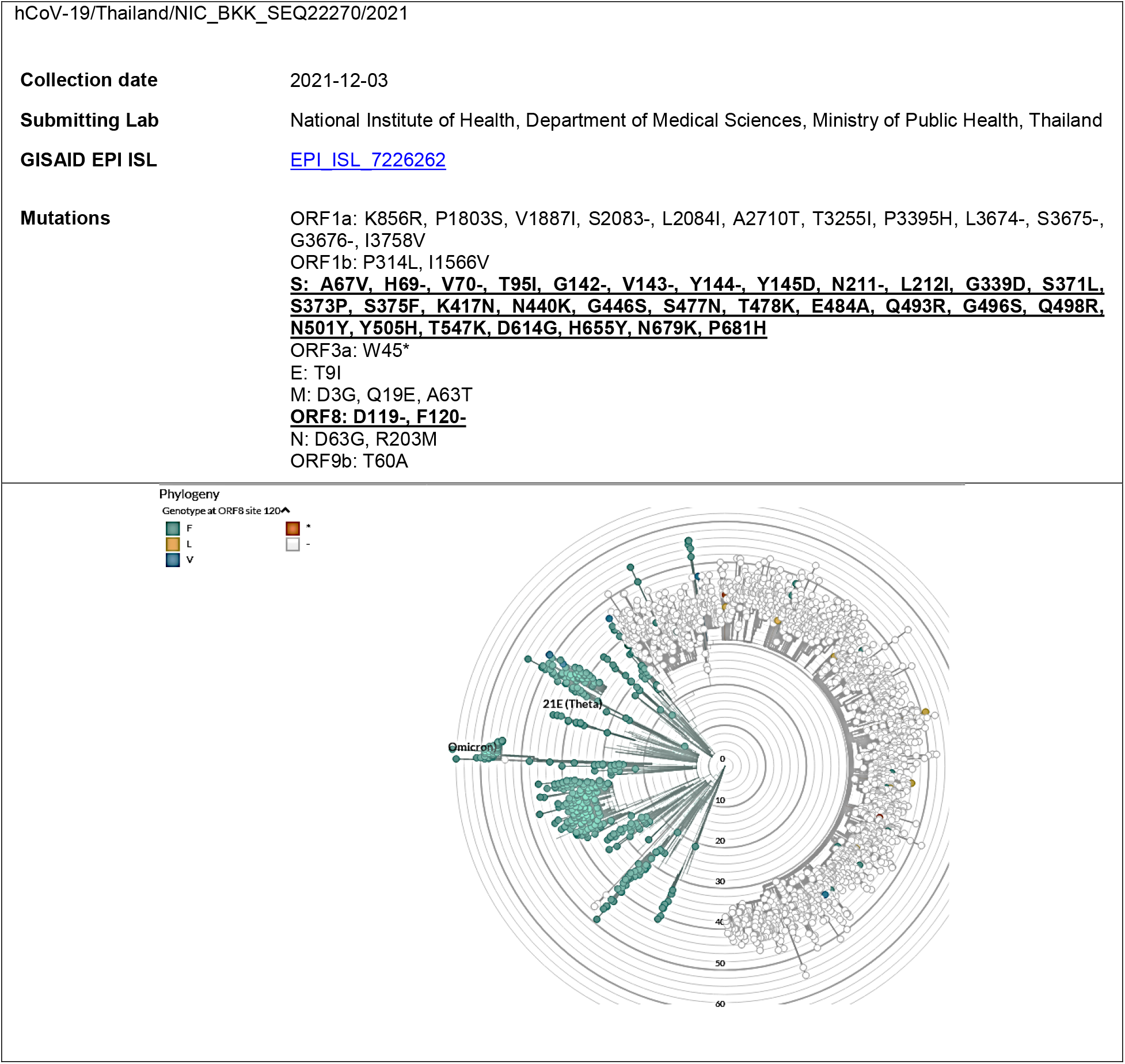
Omicron variant with the F120-mutation in ORF8 is indicated by white open circle and been fund in Sri Lanka, Thailand, Botswana, South Africa, and Ghana

## DISCUSSION

The global analysis of SARS-CoV-2 through evaluation of high entropy residues is suggestive of stochastic evolution. The original SARS-CoV-2 mutated rapidly into Alpha, Beta, Gamma, and Delta variants. The Delta variant came to dominate the pandemic and with mutational changes across the 30 high entropy residues. We are seeing mutational changes among the Delta backbone with potential of bringing about the next stochastic changes. The mutations arise in waves. The first wave consists of the structural proteins (S, M, N)- all are important in the infectivity of the virus inside the body. The second wave consists of the nonstructural ORFs which are important in the contagion of the virus outside the body ie. productive symptoms optimized to produce contagious aerosols. It is possible that being able to bind the ACE2 receptor is an important initial step in establishing infectivity follow by the ability to cause productive symptoms that can spread the virus. Infectivity serves as an anchor but not sufficient for the growth of the virus. Contagion via productive sneeze/cough is necessary to propagate the viral spread among the populace. Improving contagion before improving infectivity would fail explaining the observed mutational waves. Once both infectivity and contagion been optimized the variant-now known as Delta variant-exploded.

Most importantly are the changes affecting various ORFs. The ORF8 interactome showed a pathway cluster involving complement and coagulation cascades and cardiovascular pathology (FDR < 0.3%) [5]. Binding of ORF3a and ORF8 to TGF-β-associated factors (TGFB1, TGFB2, LTBP1, TGFBR2, FURIN, BAMBI) supports a strong involvement toward a pro-inflammation state. Moreover, a multi-omics analysis highlighted Integrin-TGF-β-EGFR-RTK signaling perturbation by ORF8. Disbalanced TGF-β signaling has been linked to lung fibrosis and oedema, a common complication of severe pulmonary diseases including COVID-19 [6]. Additionally, immune evasion is one of the unique characteristics of SARS-CoV-2 and is attributed to the ORF8 protein. This protein modulates the adaptive host immunity through downregulation of MHC-1 (Major Histocompatibility Complex) molecules and innate immune responses by suppressing the host’s interferon-mediated antiviral response. [12]

The accessory protein ORF8 is one of the most rapidly evolving betacoronavirus proteins. While ORF8 expression is not strictly essential for SARS-CoV and SARS-CoV-2 replication, a 29-nucleotide deletion (Δ29) that occurred early in human-to-human transmission of SARS-CoV, splitting ORF8 into ORF8a and ORF8b, is correlated with milder disease [7]. A 382-nucleotide deletion (Δ382) in SARS-CoV-2 [8,9] was also found to correlate with milder disease and a lower incidence of hypoxia [10]. These data would suggest that ORF8 is important in the virulence of the virus.

The crystal structure of SARS-CoV-2 ORF8 was determined at 2.04-Å resolution by X-ray crystallography. The structure reveals a ∼60-residue core similar to SARS-CoV-2 ORF7a, with the addition of two dimerization interfaces unique to SARS-CoV-2 ORF8. The last three residues _118_DFI_120_ are part of the C-terminal covalent dimer interface and could be important in the dimerization of ORF8. The dimerization of ORF8 is a relatively new evolutionary event unique to bat and human SARS-COV-2 but not SARS-CoV. [11]. Our analyses here is indicative that mutational changes to ORF8 affecting its dimerization site have a favorable evolutionary advantage to SARS-CoV-2, giving rise to the Delta variant. Additionally, mutations that are starting new clades are all within the dimerization sites. It would suggest that ORF8 un-dimerized is more capable of inducing contagion.

The Contagion Airborne Transmission inequality can be used to calculate the risk of airborne transmission of respiratory infections. Transmission occurs if extrinsic factor (Ex) is greater than intrinsic factor (N_C19_) or the droplets expelled per second times the average number of virus particles per droplet, times the fraction of droplets that make it past the face mask, times the fraction of droplets that aerosolize, times the fraction of aerosolized droplets that reach another person, times the fraction of those droplets that contain virus, times the fraction of droplets inhaled by someone not wearing a mask, times the fraction of droplets that make it through another person’s mask, times the duration of exposure is greater than or equal to the minimum inhaled viral load required to cause infection (N_C19_). N_C19_ is a physiological intrinsic value controlled by the affinity of Spike protein for ACE2 receptor and physiological stability of the virus particle in circulation as dictated by its M and N proteins. These variables occurred in the body as the virus evolved and probably evolved the fastest in immunocompromised patients such cancer patients and HIV patients. The extrinsic factor (Ex) is dependent on variables related to disease state of the host (droplet characteristics such as viral load, size, number, and percentage aerosolized and passing through mask), the environmental conditions, and the susceptible host condition. By modifying disease states the virus will be able to adapt to overcome environmental barrier such as masking, distancing, and vaccination. This is accomplished through modifications of the ORFs.

Delta variant evolved to surpass the endogenous resistant to the virus in India, it is likely that the next evolutionary offshoot from the Omicron will evolved to surpass the endogenous resistant to the virus in Africa. Further mutational changes to ORF8 could create a stronger variant of the Delta lineage or grafting of the F120-mutation onto other backbones such as the Omicron backbone could also potentially create a stronger variant. The impact on population with natural resistance could be severe but containable. However, outside of country of origin, it does raise question whether the vaccination program only against S protein will be adequate. With the number of projected infected moving to billions globally with the omicron variant, the inconceivable could be happening right now ie. the neutralizing antibodies promote instead of inhibiting viral uptake. It is imperative that we leverage our current breathing room to engineer therapeutics on top of vaccines for a long-drawn-out war against this virus.

ORF8 protein is abundantly secreted as a glycoprotein *in vitro* and in patients with newly diagnosed SARS-CoV-2. The levels of ORF8 protein in the blood correlate with disease mortality in patients with acute infection, and fatality in hospital patients is associated with higher serum levels of ORF8. Glycosylated ORF8 stimulates PBMCs to produce SARS-CoV-2 specific (IL1b, IL6, IL8) cytokines but not IL2. ORF8 induces proinflammatory cytokines through activation of NLRP3-mediated inflammasome pathways. [13]

Dysregulation of the interferon response is a strategy employed by viruses to evade host immunity. Previous investigations of the host response to SARS-CoV and MERS-CoV infection suggest that multiple coronaviruses employ this strategy [14–18]. NSP1, NSP6, NSP13 and other viral proteins interfere with IFN-beta translation [19] [20] [21]. ORF3b and ORF9b both specifically target a mitochondrial antiviral-signaling protein complex in order to inhibit type I IFN signaling [22,23]. Host factors affecting SARS-CoV-2 infection outcomes include age and sex; furthermore, comorbidities have also been implicated in interferon dysregulation [24].

SARS-CoV-2 interaction with TGF-β also caused dysregulation of NK functions [25]. Although TGF-β is thought to be an suppressed excessive immune response to the virus and its suppression would be detrimental to COVID-19 patients [26], we have proven this to be incorrect and that suppression of TGF-beta is a therapeutic option against the virus and it has been demonstrated that an untimely early production of TGF-β and associated NK cell dysfunction is a hallmark of severe SARS-CoV-2. ORF-8 has been reported to bind to and disrupt TGF-β signaling [6] as well as impairing B-cell responses against COVID-19 [27]. TGF-beta inhibitors such as OT-101, artemisinin, and prinomastat/marimastat, strongly inhibited the replication of SARS-CoV-2 but not of SARS-CoV [6].

We previously have hypothesized that TGF-β surge is the cause of SARS-CoV-2 symptoms [32–34]. Our TGB-β antisense molecule, OT-101, has completed phase 2 clinical trial against hospitalized SARS-CoV-2 patients with promising efficacy data. The phase 2 will be expanded into phase 3 at a future date. At the same time, Artemisinin, a known inhibitor of TGF-β that is also readily available as herbal supplement, was tested in our ARTI-19 clinical trial in India with similarly good clinical efficacy. We reported the safety and efficacy of Artemisinin in adult SARS-CoV-2 patients with symptomatic mild-moderate SARS-CoV-2, who were treated in a randomized, open-label Phase IV study in Bangalore, Karnataka, India (Clinical Trials Registry India identifier: CTRI/2020/09/028044). ARTIVeda showed a very favorable safety profile, and the only Artemisinin-related adverse events were transient mild rash and mild hypertension. Notably, Artemisinin, when added to the SOC, accelerated the recovery of patients with mild-moderate SARS-CoV-2. While all patients were symptomatic at baseline (WHO score = 2-4), 31 of 39 (79.5%) patients treated with Artemisinin plus SOC became asymptomatic (WHO score = 1) by the end of the 5-day therapy, including 10 of 10 patients with severe dry cough and 7 of 7 patients with severe fever. By comparison, 12 of 21 control patients (57.1%) treated with SOC alone became asymptomatic on day 5 (P=0.028, Fisher’s exact test). This clinical benefit was particularly evident when the treatment outcomes of hospitalized SARS-CoV-2 patients (WHO score = 4) treated with SOC alone versus SOC plus Artemisinin were compared. The median time to becoming asymptomatic was only 5 days for the SOC plus Artemisinin group (N=18) but 14 days for the SOC alone group (N=10) (P=0.004, Log-rank test). These data provide clinical proof of concept that targeting the TGF-β pathway with Artemisinin may contribute to a faster recovery of patients with mild-moderate SARS-CoV-2 when administered early in the course of their disease [35].

## Data Availability

All data produced in the present study are available upon reasonable request to the authors

## ACKNOWLEDGEMENT

We thank the following individuals at Brush and Key Foundation for their help in this research: Nikkita Mehta, Jeffrey Park, Andrew Ionescu, Lily Asgari.

